# Liability-scale heritability estimation for biobank studies of low prevalence disease

**DOI:** 10.1101/2022.02.02.22270229

**Authors:** Sven E. Ojavee, Zoltan Kutalik, Matthew R. Robinson

## Abstract

Theory for liability-scale models of the underlying genetic basis of complex disease provides an important way to interpret, compare and understand results generated from biological studies. In particular, liability models facilitate an understanding and comparison of the relative importance of genetic and environmental risk factors that shape different clinically important disease outcomes, through estimation of the liability-scale heritability (LSH). Increasingly, large-scale biobank studies that link genetic information to electronic health records are becoming available, containing hundreds of disease diagnosis indicators that mostly occur infrequently within the sample. Here, we propose an extension of the existing liability-scale model theory suitable for estimating LSH in biobank studies of low-prevalence disease. In a simulation study, we find that our derived expression yields lower MSE and is less sensitive to prevalence misspecification as compared to previous transformations, for diseases with ≤ 2% population prevalence and LSH of ≤ 0.45, especially if the biobank sample prevalence is less than that of the wider population. Applying our expression to 13 diagnostic outcomes of ≤ 3% prevalence in the UK Biobank study, revealed important differences in LSH obtained from the different theoretical expressions, that impact the conclusions made when comparing LSH across disease outcomes. This demonstrates the importance of careful consideration for estimation and prediction of low prevalence disease outcomes, and facilitates improved inference of the underlying genetic basis of ≤ 2% population prevalence diseases, especially where biobanking sample ascertainment results in a healthier sample population.

## Introduction

Genetically informed deep-phenotyped biobanks are an increasingly available important research resource. From these data, estimates of SNP heritability,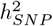, can be obtained, a quantity describing the proportion of phenotypic variance attributable to the genetic marker data [1]. Linked electronic health records, provide a large number of binary, presence/absence disease diagnosis indicators and it is important to be able to compare 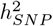 estimates, in order to infer the relative importance of genetic and environmental risk factors that shape different clinically important disease outcomes.

To better describe the genetics of such binary traits, the notion of liability scale heritability (LSH) has been coined [2] to reflect the underlying continuous nature of additive genetic effects. An initial derivation for LSH was given by Alan Robertson in the Appendix of Dempster and Lerner [3] for the scenario where the case-control ratio was the same in the sample and the population. Lee et al. [4], proposed an extended derivation to account for the fact that, in a case-control study, cases tend to be over-represented compared to the population prevalence, arriving at the following expression for the LSH

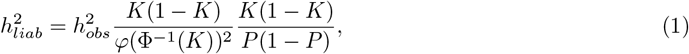

where, 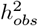 is the observed scale heritability, *K* is the prevalence of the binary trait in the full population, *P* is the prevalence of the binary trait in the sampled subpopulation, and the denominator of the first fraction is the squared probability density function of the standard normal distribution evaluated at the *K*th quantile of the inverse cumulative density function of the standard normal distribution. This expression was derived under the assumption that sample prevalence is greater than or equal to population prevalence (*P* ≥ *K*).

The problem of accurately estimating LSH was further investigated by Golan et al. [5] who noted that in the common setting of sample prevalence exceeding population prevalence (*P > K*), Eq.1 applied on REML estimates underestimates LSH. To account for this, they proposed phenotype correlation–genotype correlation (PCGC) regression, which generalises a Haseman–Elston regression and yields unbiased estimates of LSH in simulations. In many settings, especially if there are individual-level data available and the sole goal of the analysis is to estimate the LSH, it is recommended to use PCGC. However, often only summary-level marginal SNP regression coefficients are provided by biobank studies, meaning that 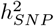 must be indirectly inferred (e.g. using LD score regression [6] or High-definition likelihood [7]), or observed scale heritability estimate is a hyper-parameter embedded into effect size estimation (BayesRR-RC [8]). Therefore, it remains important to facilitate the transformation of observed scaled heritability into LSH.

The structure of the biobank data sets creates additional problems. Firstly, they often represent a subset of the population that often is healthier, younger, or has higher socio-economic background than the average population. Because of this, many if not most binary disease traits have a lower sample prevalence compared to the population prevalence (*P < K*). The classical expression in Eq.1 has been derived and tested for situations where *P* ≥ *K*, and this can result in estimates with inflated variance for situations where *P < K*. Secondly, the disease prevalence in biobank-scale studies can be small and measurement errors of such a small quantity could greatly amplify the variance of the LSH estimate. In practice, as biobanks include people born during different decades, cohorts have usually not reached the end of their lifespan and as disease prevalences change across time, it seems appropriate to accompany the prevalence estimates with error estimates when arriving at an LSH estimate. Furthermore, although Eq.1 has solid theoretical justification, it does not guarantee that the LSH estimate will be lower than or equal to 1, and even if the LSH estimate is bounded, it can still have high variance in the biobank setting, especially if the model is imperfectly specified or there are greater deviations away from the assumption of the existence of latent genetic liability.

Here, we propose an alternative expression to address some of those issues. Firstly, the suggested expression will guarantee that the LSH estimate will be bounded between 0 and 1, and secondly, we demonstrate that for low prevalences (*K* ≤ 0.02) our formula results in lower MSE compared to the classical expression. Thirdly, we show that our formula limits the inflation in MSE if we take into account the uncertainty in the prevalence estimation. Although our expression is also derived under the simplifying assumption of *P* = *K*, we argue that for many biobank-based studies this assumption is preferred as long as *P* ≤ *K* and the prevalence is small. Finally, we apply our proposed expression on 13 disease outcomes with low sample prevalence in the UK Biobank and we compare the LSH estimates obtained by our expression to those of Eq.1. We also provide a shiny app https://medical-genomics-group.shinyapps.io/h2liab/.

### Derivation of the expression

Suppose that we have a binary trait with a frequency of *K* in the population and a frequency of *P* in the sample is here equal to *K*. Suppose that a liability model holds meaning that there exists a latent liability *l* that is defined as a sum of genetic (*g*) and error (*e*) components *l* = *g* + *e*. We assume that *l* has a variance of 1 and that *g* and *e* come from normal distributions: 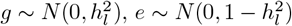, where 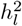 is the LSH. Then we assume that the binary disease trait *y* is associated with *l* such that *y* = 1 if *l > t* and *y* = 0 if ≤ *t*, where *t* is some liability scale threshold defining the required liability value for disease occurrence, and *t* = Φ^−1^(1 − *K*).

We first write the expression how is the observed scale heritability associated with LSH. As shown by Dempster and Lerner [3]

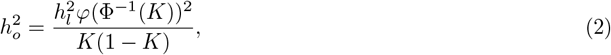

where 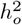 is the observed scale heritability. We recognise that the numerator represents the observed scale genetic variance and the denominator represents total observed scale phenotypic variance. Our idea is to replace the total phenotypic variance estimate *K*(1 − *K*) with the sum of genetic and error variances and that by definition will guarantee that the LSH estimate remains bounded. Thus, we need to rewrite the total phenotypic variance using the error variance *E*(*V ar*(*y*|*c* + *zg*))

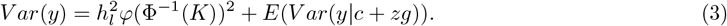

We find (see Supplementary Information) that the error variance is expressed as

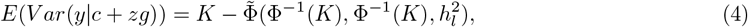

where 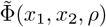 is the cumulative distribution function of a bivariate normal distribution with a mean of 0, variance of 1 and correlation *ρ* evaluated at (*x*_1_, *x*_2_). That gives us a the expression for observed scale heritability

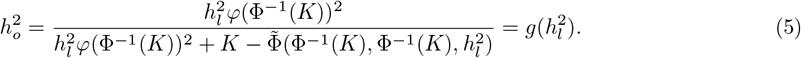

It is impossible to find the closed expression for 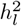 from the last expression, however, we can still plug in values for *K* and 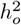 and solve Eq.5 numerically for 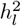.

We derive the variance for 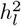 from the last expression using the delta method. Suppose that we know 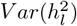 and we would like to find 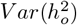. For this, we need to differentiate expression 5 with respect to 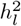 and that results in

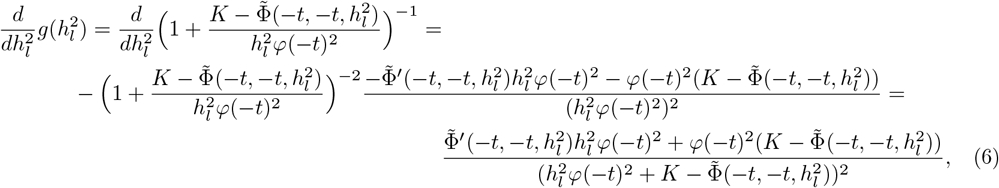

where we have denoted Φ^−1^(*K*) = −*t* (negative of the disease defining threshold), 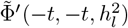 is the partial derivative of 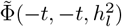 with respect to 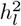 and as demonstrated by Drezner and Wesolowsky [9]

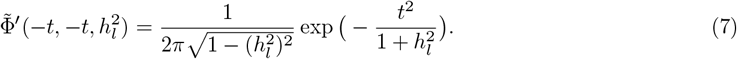

The variance can thus be expressed from the delta method as

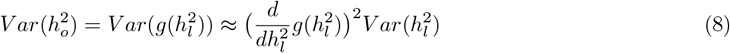

and conversely for the 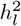

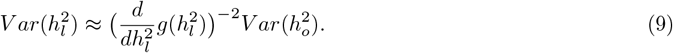

### Simulation study

We executed two simulation settings. Simulation 1 followed the strategy of Lee et al. [4] and we used it to demonstrate the implications of the uncertainty of prevalence estimates to the outcome in the absence of ascertainment. There, we used a sample size of 20,000 and we created a low level of relatedness by simulating individuals in independent batches of 100 with genetic values (*g*) drawn from a multivariate normal distribution given a 100 × 100 covariance matrix with off-diagonal elements in the covariance matrix were 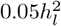 and 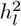 for the diagonals. Error term *e* was simulated from a normal distribution 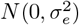 such that the variance of the liability *l* = *g* + *e* would be 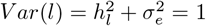. The corresponding observed binary phenotype was defined as

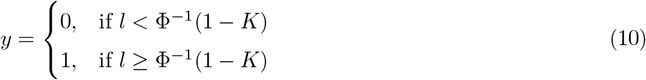

We varied the true LSH from 0.15 to 0.65 with a step of 0.1 and we varied prevalence between *K* =0.001, 0.005, 0.01, 0.05, 0.1, 0.5. In addition, to mimic the potential uncertainty coming from the heterogeneity across estimates, at each step when estimating the LSH, we drew the value for 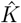 from a normal distribution *N*(*K*, (0.1*K*)^2^) yielding a coefficient of variation of 10%. The number of simulation replicates was 2000.

For low prevalence traits, our proposed formula results in a more accurate result in terms of MSE in many common settings where the LSH is smaller than 0.55 and the prevalence less than 2% (Figure 1). However, we note caution that if the underlying LSH is higher than 0.45, then bounding the LSH estimate below 1 results in a small bias that is observed as the prevalence approaches 1% or greater (Figure 1), but we expect this to be the exception for real-world disease outcomes. Using an inaccurately measured prevalence results in increased MSE, with the new proposed formula being more robust to this kind of misspecification, as it yields lower MSE increase across all scenarios (Figure 3).

**Figure 1.**
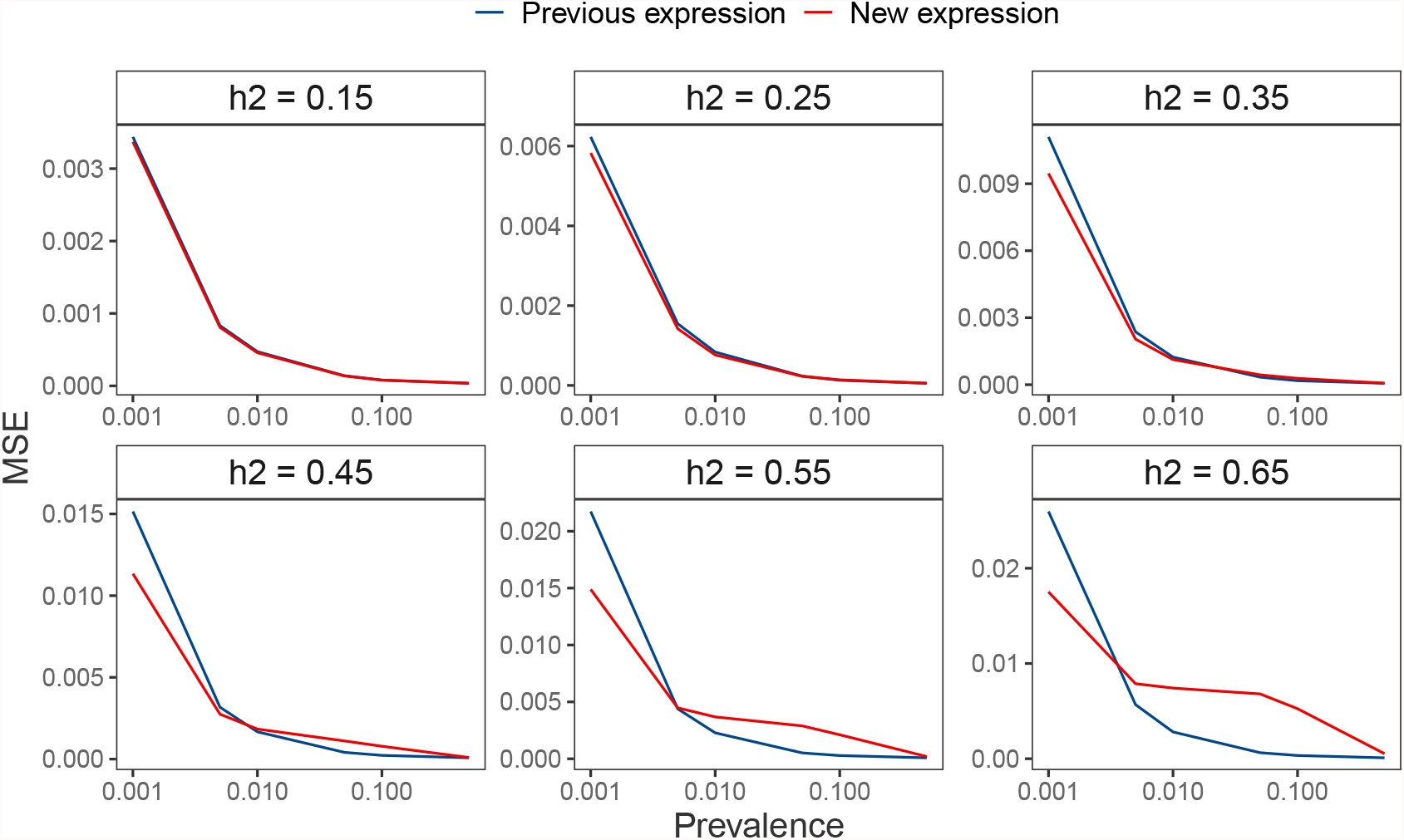
MSE of previous and new formula in the case of no ascertainment and no measurement error for the prevalence across different values for the liability-scale heritability. The new estimate yields lower MSE in the settings where the prevalence is low (*K <* 0.01). The benefit of using the new formula decreases as the underlying liability-scale heritability increases, stemming from the small bias introduced in the new estimate that is also bounding the estimate below 1. Data sets created under simulation scenario 1.

**Figure 2.**
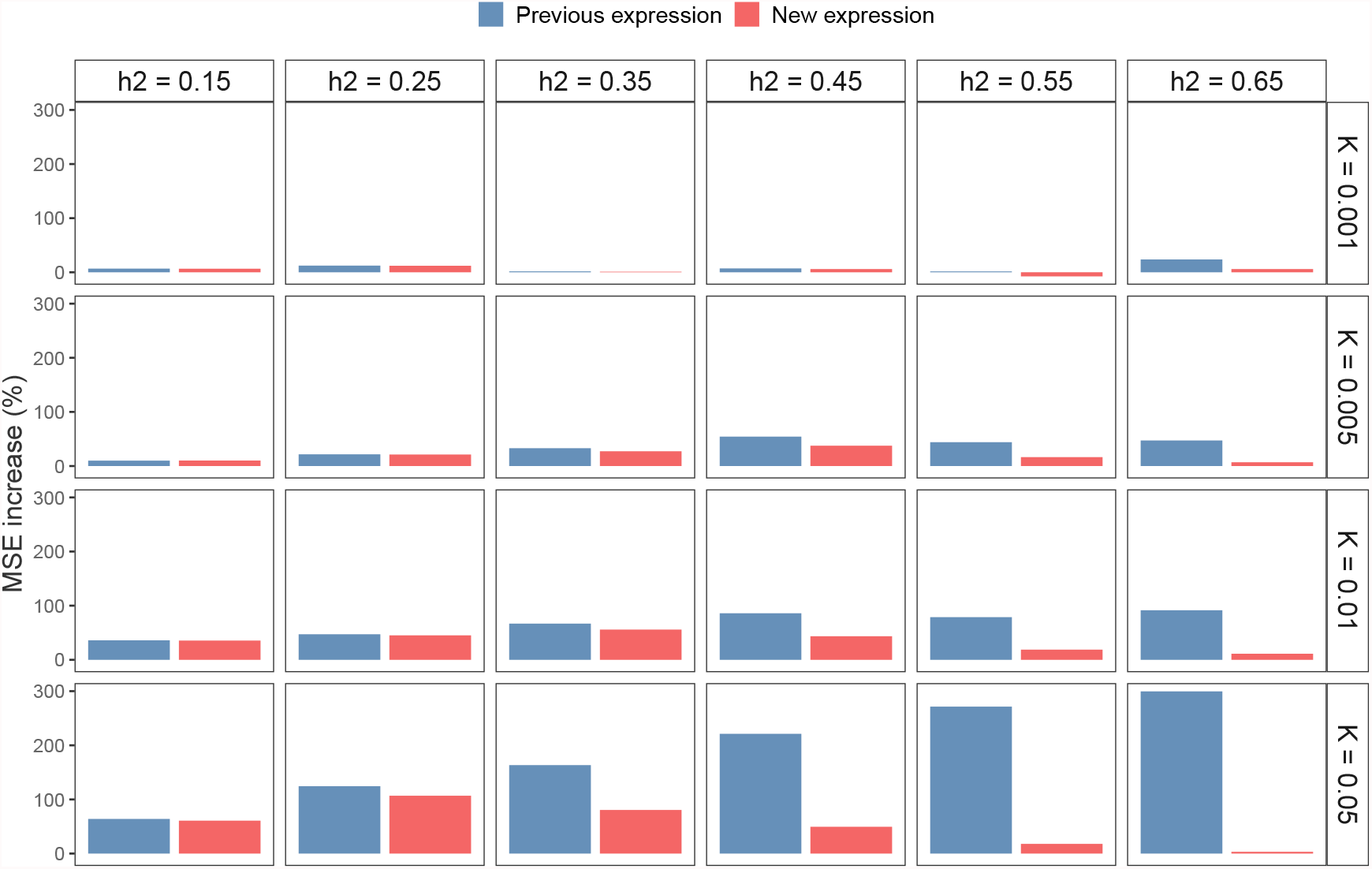
Increase in MSE due to measurement error of prevalence. Using prevalence that has not been completely accurately measured results in increase in the mean squared error. However, the new proposed formula is more robust to this kind of misspecification as it yields lower MSE increase across all scenarios. Data sets created under simulation scenario 1.

**Figure 3.**
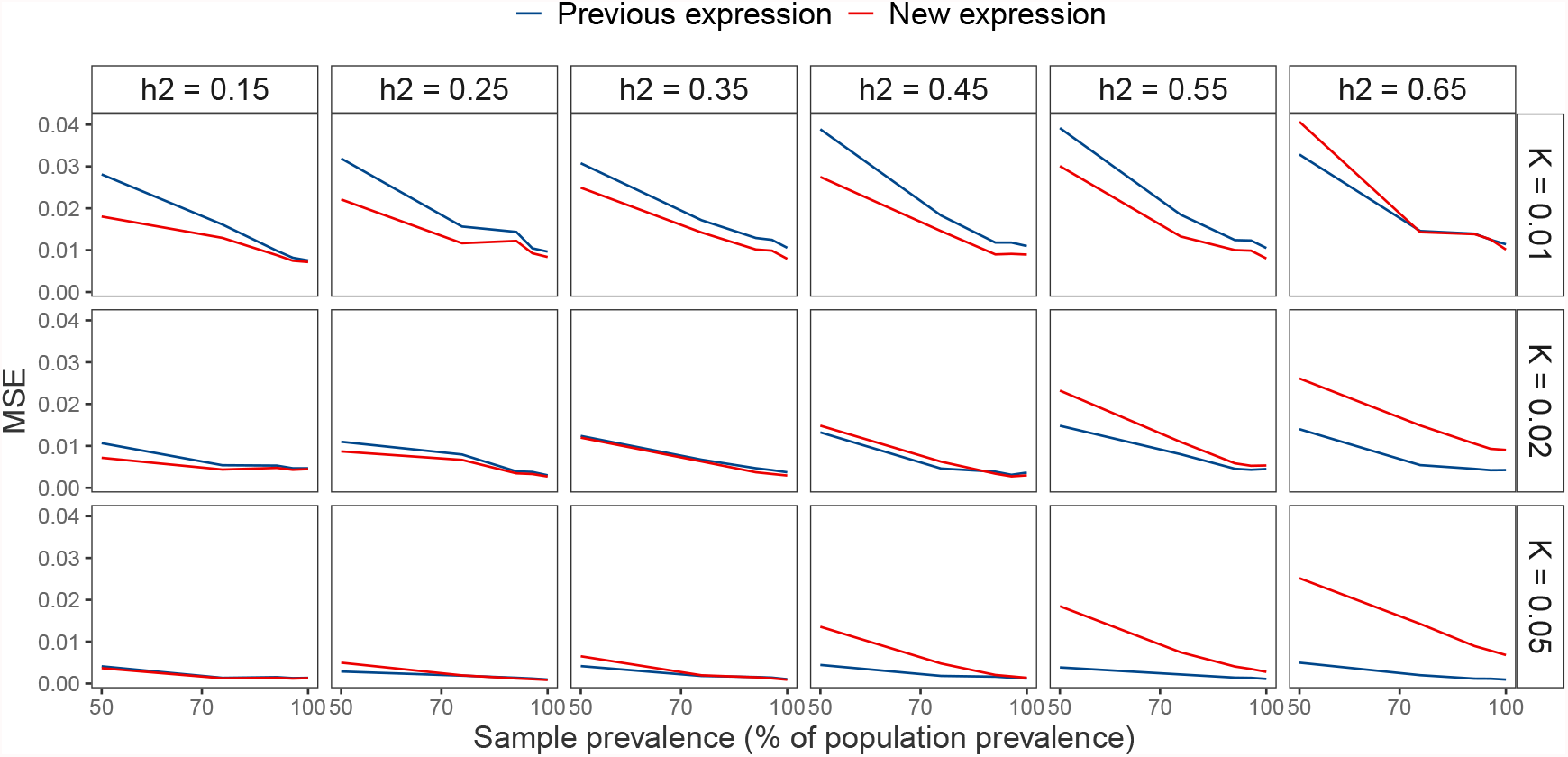
Differences in MSE if sample prevalence is lower than the population prevalence. The new expression works better at lower population prevalences (*K* ≤ 0.02) and with moderate or low LSH values 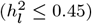. Data sets created under simulation scenario 2.

Simulation 2 examined a more realistic scenario similarly to Golan et al. [5]. The final sample size was again 20,000 but here we created the genetic values using SNP data of 10,000 markers. For each of the 10,000 SNPs, we simulated the SNP MAF from a uniform distribution *U*(0.05, 0.5) and the minor allele counts *x*_*ij*_ for individual *i* at SNP *j* were simulated from a binomial distribution *B*(2,*MAF*_*j*_). The effect size for each SNP *j* was drawn from a normal distribution such that 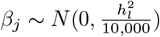. The genetic value for an individual *i g*_*i*_ was calculated as 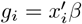 and the error term *e*_*i*_ was simulated from 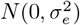 such that the variance of the liability *l*_*i*_ = *g*_*i*_ + *e*_*i*_ would be 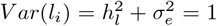. The liability scale phenotype *l*_*i*_ was translated into binary phenotype *y*_*i*_ as shown in Eq.10. To create suitably ascertained samples we first simulated a slightly larger population of *Ñ*(*>*20,000) that has a case prevalence (in population) of *K* and then we randomly selected *N* · *P* cases and *N* · (1 − *P*) controls to achieve case prevalence (in sample) of *P*. We used GREML [10] from GCTA [11] to estimate the observed scale heritability. Finally, we used the Eq.1 to transform it to the known LSH estimate and we used Eq.5 by using *P* as the prevalence to yield the new LSH estimate. We used the same values for heritability, we varied the *K* between 0.01, 0.02 and 0.05, and we varied *P* between 50%, 75%, 90%, 95%, 100% of the *K* value. Due to the increased computational complexity, we resorted to 100 simulation replicates.

Here, we find the new expression of Eq.5 gives lower MSE at lower population prevalence (*K* ≤ 0.02) and with moderate or low LSH values (LSH ≤ 0.45) as compared to the classical expression of Eq.1, supporting the results of the first simulation setting (Figure 4). Therefore, we propose that for biobank studies of diseases with ≤ 2% population prevalence, especially where the sample prevalence is less than that of the wider population, Eq.5 should be preferred over Eq.1.

**Figure 4.**
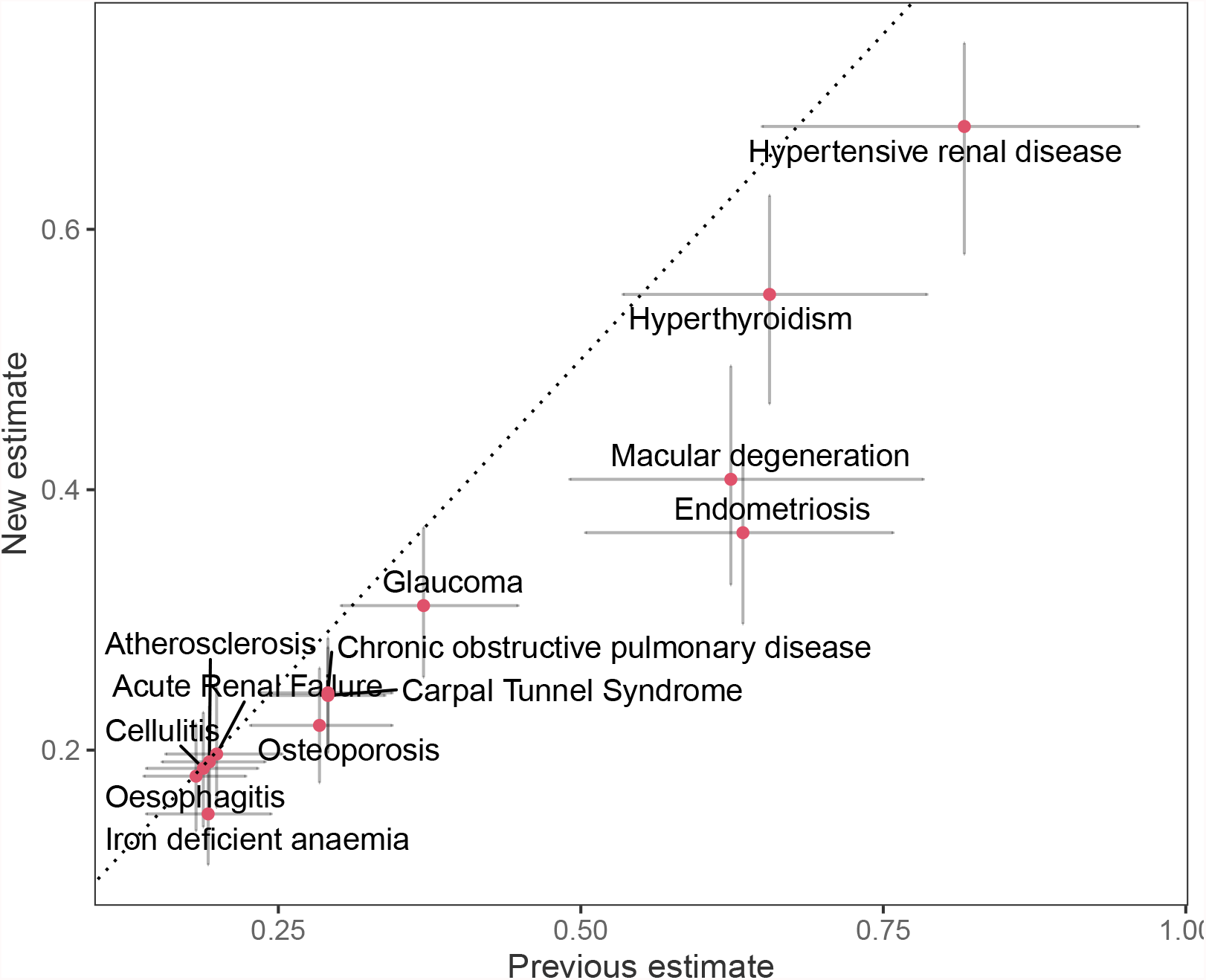
Comparison of previous and new liability-scale heritability estimates with 95% CI for 13 ICD-10 binary disease outcomes with ≤ 3% sample prevalence in the UK Biobank. Table 1 gives the full disease names, codes, sample prevalence, population prevalence, observed-scale estimates, and liability-scale conversions. We analysed the 13 traits with our recently proposed GMRM software, to obtain observed scale heritability estimates, accounting for marker effect size differences across SNPs of different minor allele frequency, linkage disequilibrium and functional annotation in 382,390 unrelated UK Biobank individuals. We estimated the posterior mean observed scale heritability and plot the liability scale estimates produced with either using the classical expression of Eq.1 on the x-axis or our new proposed expression of Eq.5 on the y-axis.

### Empirical data analysis

We then analysed 13 ICD-10 binary disease outcomes with ≤ 3% sample prevalence in UK Biobank (Table 1) with our recently proposed GMRM software, to obtain observed scale heritability estimates, accounting for marker effect size differences across SNPs of different minor allele frequencies, linkage disequilibrium and functional annotation [8, 12], using 2,174,071 SNP markers and 382,390 individuals of SNP marker relatedness (≤ 0.05). To attempt to control for environmental confounding before analysis, we adjusted for the leading 20 principle components as supplied by the UK Biobank, sex as a binary factor, age as a linear and quadratic term, east and north coordinate of residence, recruitment centre, and genotype batch. We estimated the posterior mean observed scale heritability from the last 1,500 sampling iterations after stabilisation of the running mean, and then compared the liability scale estimates produced with either using the classical expression of Eq.1 or our new proposed expression of Eq.5.

**Table 1.**
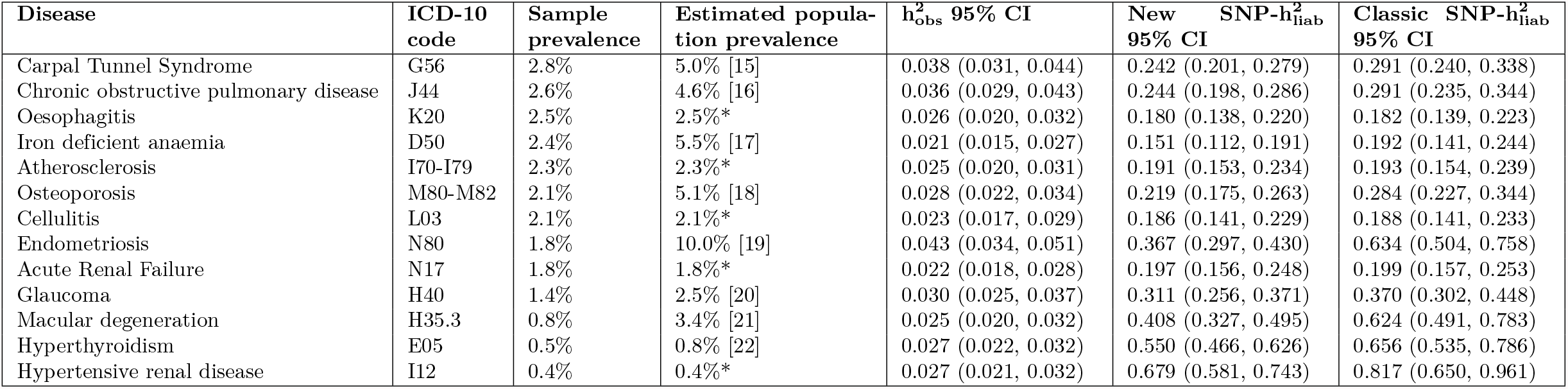
13 selected disease outcomes recorded in the UK Biobank of <3% sample prevalence with higher prevelence in the general UK population. Columns of the table give the commonly used disease name, the ICD-10 code, the UK Biobank sample prevalence, and the estimated UK population prevalence with corresponding reference (* denotes that we used the UK Biobank due to unavailability or vast heterogeneity in the estimates), the posterior mean 0/1 observed scale single nucleotide polymorphism (SNP) heritability with 95% credible interval.

| Disease | ICD-10 code | Sample prevalence | Estimated population prevalence | $h^2_{\text{obs}}$ 95% CI | New SNP-$h^2_{\text{liab}}$ 95% CI | Classic SNP-$h^2_{\text{liab}}$ 95% CI |
| --- | --- | --- | --- | --- | --- | --- |
| Carpal Tunnel Syndrome | G56 | 2.8% | 5.0% [15] | 0.038 (0.031, 0.044) | 0.242 (0.201, 0.279) | 0.291 (0.240, 0.338) |
| Chronic obstructive pulmonary disease | J44 | 2.6% | 4.6% [16] | 0.036 (0.029, 0.043) | 0.244 (0.198, 0.286) | 0.291 (0.235, 0.344) |
| Oesophagitis | K20 | 2.5% | 2.5%* | 0.026 (0.020, 0.032) | 0.180 (0.138, 0.220) | 0.182 (0.139, 0.223) |
| Iron deficient anaemia | D50 | 2.4% | 5.5% [17] | 0.021 (0.015, 0.027) | 0.151 (0.112, 0.191) | 0.192 (0.141, 0.244) |
| Atherosclerosis | I70-I79 | 2.3% | 2.3%* | 0.025 (0.020, 0.031) | 0.191 (0.153, 0.234) | 0.193 (0.154, 0.239) |
| Osteoporosis | M80-M82 | 2.1% | 5.1% [18] | 0.028 (0.022, 0.034) | 0.219 (0.175, 0.263) | 0.284 (0.227, 0.344) |
| Cellulitis | L03 | 2.1% | 2.1%* | 0.023 (0.017, 0.029) | 0.186 (0.141, 0.229) | 0.188 (0.141, 0.233) |
| Endometriosis | N80 | 1.8% | 10.0% [19] | 0.043 (0.034, 0.051) | 0.367 (0.297, 0.430) | 0.634 (0.504, 0.758) |
| Acute Renal Failure | N17 | 1.8% | 1.8%* | 0.022 (0.018, 0.028) | 0.197 (0.156, 0.248) | 0.199 (0.157, 0.253) |
| Glaucoma | H40 | 1.4% | 2.5% [20] | 0.030 (0.025, 0.037) | 0.311 (0.256, 0.371) | 0.370 (0.302, 0.448) |
| Macular degeneration | H35.3 | 0.8% | 3.4% [21] | 0.025 (0.020, 0.032) | 0.408 (0.327, 0.495) | 0.624 (0.491, 0.783) |
| Hyperthyroidism | E05 | 0.5% | 0.8% [22] | 0.027 (0.022, 0.032) | 0.550 (0.466, 0.626) | 0.656 (0.535, 0.786) |
| Hypertensive renal disease | I12 | 0.4% | 0.4%* | 0.027 (0.021, 0.032) | 0.679 (0.581, 0.743) | 0.817 (0.650, 0.961) |

Liability-scale heritability estimates obtained by Eq.5 were lower than the classical expression of Eq.1 (Figure 4). For disease outcomes, where we assume that the UK Biobank sample prevalence is identical to the wider population prevalence, the estimates from either equation are in agreement (Figure 4). However, once the sample prevalence was ≤ 1%, and lower than that estimated in the wider population from which it was drawn, we observe substantially lower liability-scale estimates from Eq.5 (Figure 4). For example, LSH differed by 0.26 for endometriosis and 0.22 for macular degeneration although this difference is mainly driven by the difference in population and sample prevalences. Even though we assumed equal sample and population prevalence for hypertensive renal disease, we still get a difference of 0.14 between the classical and new estimates accompanied by two times narrower 95% CI for the new estimate (Table 1). These differences influence the inference made, as Eq.5 estimates no significant difference of LSH for glaucoma, endometriosis, and macular degeneration, in contrast to the results that would be obtained from Eq.1 (Figure 4). Therefore, the lower MSE of our proposed estimate across many common settings translates to real-world differences in inference for low prevalence diseases within biobank studies.

## Discussion

Using biobank-scale data sets to infer the heritability of rare traits is increasingly important, due to the general difficulties of collecting case-control samples for rare diseases. However, our results demonstrate the importance of treating low prevalence traits with extra care. Moreover, the advantage of using our proposed estimate is even greater if we take into account the uncertainty or heterogeneity in the lifetime prevalence estimates. Heterogeneity in the lifetime prevalence estimates is very common and stems from regional differences, differences in the time of the study, methodology, usage of subpopulations, and often the lifetime prevalence estimate is simply unavailable or some proxies are used. All this amounts to a considerable amount of uncertainty and unfortunately, the most common way to handle this is to simply pick one of the prevalence estimates and not take into account the uncertainty. Our proposed estimate limits the error coming uncertainty and we argue that the future analyses using lifetime population prevalence should reflect the uncertainty in the estimates.

The analysis of empirical data in UK Biobank calculating SNP heritability demonstrated that in many settings the classical expression could lead to likely, if not clear overestimates. For example for hypertensive renal disease, the classic formula gave an LSH estimate of 0.82 (95% CI 0.65, 0.96) whereas the new formula estimated LSH of 0.68 (95% CI 0.58, 0.74). For a similar trait of chronic kidney disease, the heritability values have been estimated between 0.30 to 0.75 [13] suggesting that the classical estimate might be unrealistically high. We also observe a similar effect for macular degeneration where the classical formula also yields a likely overestimate of 0.62 (95% CI 0.49, 0.78) whereas the literature suggests SNP heritability of [14] of 0.47 that had been calculated on a larger case count.

In general, there seems to be a switch point in prevalence after which the classical expression (Eq.1) tends to become more effective. That is probably because *K*(1 − *K*) is a good estimator for the total phenotypic variance with high *K* values (as in Eq.2) but with small *K* values that product *K*(1 − *K*) becomes tiny and the expressions using the inverse 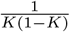 will become highly sensitive to the observed scale heritability estimation error. Our proposed expression makes the total phenotypic variance dependent on the estimated observed genetic variance and thus controls better for the mismatch between the total phenotypic and genotypic variances in the classical expression.

There are important caveats to our proposed formula. Even though we manage to effectively constrain the heritability between 0 and 1, it also introduces a small downwards bias that becomes more visible with higher values of prevalence and true liability scale heritability (>0.6). Nevertheless, we argue that this will likely be the exception for real-world disease outcomes. Any transformation of scale will be an approximation, made under a set of theoretical assumptions and our aim here is to simply provide an approach that facilitates comparisons of the proportion of variance attributable to the SNP markers for low prevalence diseases with as low MSE as possible. Secondly, our formula assumes that the biobank prevalence is approximately reflecting the population prevalence. In most low prevalence scenarios we believe this approach is justified since the sample prevalence is similar or slightly smaller compared to the population prevalence, the latter case resulting in more conservative or lower MSE estimates (Figure 4). In principle, it is possible to extend the Eq 5 to account for ascertainment. However, the outcome when expressing total phenotypic variance as a sum of error and genetic variances would still depend on products of *K*(1 − *K*) and *P*(1 − *P*) and thus this approach would not have any benefit in terms of reducing MSE.

Here, we have proposed a new expression for calculating LSH suitable for traits with low prevalence. We have shown that our proposed formula results in a more accurate LSH estimator in terms of MSE in many common settings and in general results in a slightly more conservative estimate that can result in more accurate estimates of liability scale heritability. Hopefully, it can lead to a more realistic quantification of rare trait heritabilities, many of which are still yet to be explored.

## Data Availability

All data produced in the present study are available upon reasonable request to the authors.

## Acknowledgements

This project was funded by an SNSF Eccellenza Grant to MRR (PCEGP3-181181), and by core funding from the Institute of Science and Technology Austria. This research was supported by the Scientific Service Units (SSU) of IST Austria through resources provided by Scientific Computing (SciComp).

## Author contributions

SEO and MRR conceived and designed the study. SEO derived the equations. ZK provided study oversight. SEO and MRR analysed the data and wrote the paper. All authors approved the final manuscript prior to submission.

## Supplementary Information

### Derivation of the error variance term

Suppose that the genetic value (of an individual) *g* and error term *e* are from normal distributions 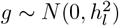 and 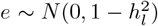, where 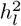 is the underlying liability scale heritability. Then the underlying liability *l* = *g* + *e* and the binary trait *y* with a prevalence of *K* is defined as

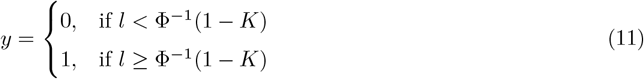

From this we will derive the error variance term *E*(*V ar*(*y*|*c* + *zg*)) where *c* is some constant and *z* is the standard Gaussian density evaluated at Φ^−1^(1 − *K*) as shown in [3]. As *c* + *zg* is a linear combination of *g* we can equivalently find *E*(*V ar*(*y*|*g*)). First, we note the conditional distribution of *y* given *g*

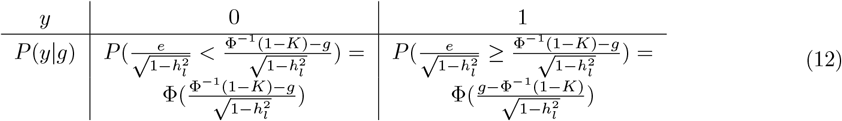

As *y* can be equal to only 0 or 1, we can write

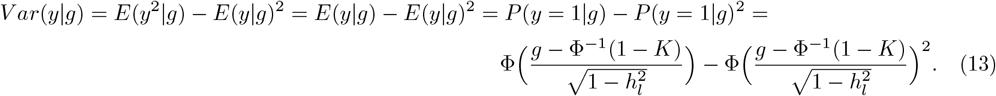

To find *E*(*V ar*(*y*|*g*)) we need to find 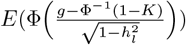 and 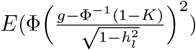. For this, we use auxiliary standardised Gaussian random variables *X, X*_1_ and *X*_2_ that are independent of *g* and *X*_1_ is independent of *X*_2_. From this it follows that 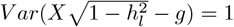 and using the law of total probability we get

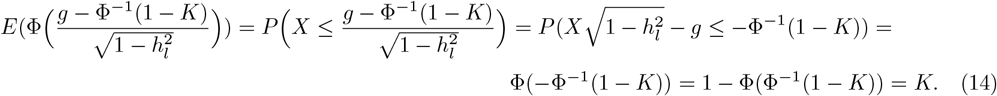

Secondly, we see that we can analogously use *X*_1_ and *X*_2_ to find the second moment of 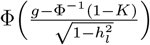. For this we need to find the following correlation

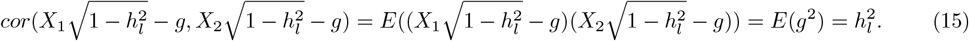

Now we express the expectation using a cumulative distribution function of a bivariate Gaussian distribution of two random variables that have a correlation of 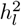

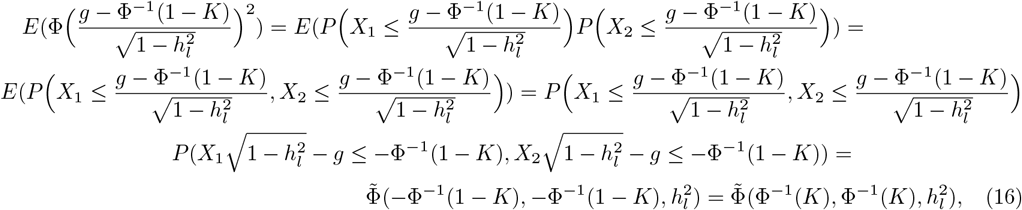

where 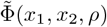 is the cumulative distribution function of a standardised bivariate Gaussian distribution with a correlation of *ρ*. The first equation follows from the definition of cumulative distribution function, second from the independence of *X*_1_ and *X*_2_, third from the law of total probability. Thus, by combining the two last results, we get the final expression for the error variance

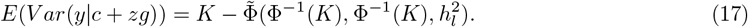

